# Vaccine Effectiveness against COVID-19 among Symptomatic Persons Aged ≥12 Years with Reported Contact with COVID-19 Cases, February – September 2021

**DOI:** 10.1101/2021.12.30.21267928

**Authors:** Jessie R Chung, Sara S Kim, Edward A Belongia, Huong Q McLean, Jennifer P King, Mary Patricia Nowalk, Richard K Zimmerman, Krissy Moehling Geffel, Emily T Martin, Arnold S Monto, Lois E Lamerato, Manjusha Gaglani, Eric Hoffman, Marcus Volz, Michael L Jackson, Lisa A Jackson, Manish M Patel, Brendan Flannery

**Affiliations:** Centers for Disease Control and Prevention, Atlanta, GA, USA; Marshfield Clinic Research Institute, Marshfield, WI, USA; University of Pittsburgh Schools of the Health Sciences and University of Pittsburgh Medical Center, Pittsburgh, PA, USA; University of Michigan, Ann Arbor, MI, USA; Henry Ford Health System, Detroit, MI, USA; Baylor Scott and White Health; Texas A&M University College of Medicine, Temple, TX, USA; Kaiser Permanente Washington Health Research Institute, Seattle, WA, USA

**Keywords:** SARS-CoV-2, COVID-19, Vaccine Effectiveness

## Abstract

Individuals in contact with persons with COVID-19 are at high risk of developing COVID-19, but protection offered by COVID-19 vaccines in the context of known exposure is unknown. Symptomatic outpatients reporting acute onset of COVID-19-like illness and tested for SARS-CoV-2 infection were enrolled. Among 2,229 participants, 283/451 (63%) of those reporting contact and 331/1778 (19%) without known contact tested SARS-CoV-2 positive. Using the test-negative design, adjusted vaccine effectiveness was 71% (95% confidence interval, 49%-83%) among fully vaccinated participants reporting contact versus 80% (95% CI, 72%-86%) among those without. This study supports COVID-19 vaccination and highlights the importance of efforts to increase vaccination coverage.

## Introduction

Individuals in contact with persons with COVID-19 disease are at high risk of SARS-CoV-2 infection and developing COVID-19 disease themselves [1]. The US Centers for Disease Control and Prevention (CDC) recommends vaccination as the best tool to prevent COVID-19 disease among persons aged ≥5 years in conjunction with non-pharmaceutical interventions (NPIs) such as hand washing, mask wearing, and physical distancing [2]. Repeated, extended exposures in close proximity to persons with SARS-CoV-2 infection can increase the risk of becoming infected [3]. Having close contact with a person with COVID-19 disease, such as within a household, is one of the main sources of new SARS-CoV-2 infections. Thus, CDC recommends persons who are not yet fully vaccinated to seek testing immediately after finding out they have had close contact, even if they do not have symptoms [4].

Data are limited regarding how COVID-19 vaccine effectiveness (VE) may vary by the intensity of the exposure [5]. One approach to assess the impact of exposure would be to evaluate VE against symptomatic illness among individuals with known contact, compared to individuals unaware of close exposure. These data may contribute to efforts to increase COVID-19 vaccine uptake among persons who have not yet received vaccines, and possibly inform NPI strategies as coverage increases [5]. In this report, we build upon prior studies of COVID-19 VE from the US Influenza Vaccine Effectiveness (Flu VE) Network to present VE against symptomatic laboratory-confirmed SARS-CoV-2 infection among persons aged ≥12 years with and without known contact with a person with SARS-CoV-2 infection from February through September 2021.

## Methods

Methods used for estimating VE against laboratory-confirmed symptomatic COVID-19 disease among persons seeking medical care or testing for SARS-COV-2 infection in Flu VE Network study sites have been previously described [6]. Briefly, research staff screened persons who sought outpatient medical care (i.e., telehealth, primary care, urgent care, and/or emergency department) or clinical SARS-CoV-2 testing using a standard case-definition for COVID-like illness that included acute onset of fever/feverishness, cough, or loss of taste or smell, with symptom duration <10 days [7]. Research staff contacted potentially eligible outpatients either in person, by telephone, or email to confirm eligibility and enroll participants who consented verbally or in writing. Standardized questionnaires collected demographic information, self-reported contact ≤14 days before illness onset with a person with confirmed COVID-19 (i.e., “known contact”), healthcare-related occupation, and COVID-19 vaccination. Participants had SARS-CoV-2 molecular testing on respiratory specimens collected within 10 days of illness onset; results were used to classify SARS-CoV-2-positive cases and test-negative controls.

For this analysis, we included participants aged ≥12 years with illness onset between February 1 and September 30, 2021. Beginning dates of inclusion in analyses varied by site according to local COVID-19 vaccination policies for all persons by age group as vaccines became available (February 1, 2021 for persons aged ≥65 years, March 22, 2021 for persons aged 16–64 years, and May 12, 2021 for persons aged 12–15 years). We determined vaccination status through participant interviews, and verified vaccination based on participant-provided vaccination record cards, and/or documentation of vaccination in electronic medical or state immunization information systems. Fully vaccinated participants were defined as those who received two doses of an mRNA vaccine (Pfizer-BioNTech BNT162b2 or Moderna mRNA-1273) or one dose of Johnson and Johnson/Janssen vaccine (JNJ-784367350) ≥14 days before illness onset [8]. Partially vaccinated participants were defined as those who received at least one dose of an mRNA vaccine ≥14 days before illness onset but who were not fully vaccinated. Those who had no documentation of any COVID-19 vaccination prior to illness onset were defined as unvaccinated. Participants whose first documented dose was received <14 days prior to illness onset (n=141) were excluded from VE analyses.

We used the test-negative design to evaluate VE of currently available SARS-CoV-2 vaccines against symptomatic, laboratory-confirmed outpatient COVID-19 disease [9] among participants reporting contact with persons with confirmed COVID-19 disease and those reporting no known contact. Persons who did not complete the question regarding known contact were excluded from primary analyses. VE was calculated as 1 – adjusted odds ratio of vaccination among symptomatic SARS-CoV-2-test-positive participants versus symptomatic test-negative participants (controls) using multivariable logistic regression. An interaction term for known contact and vaccination status was assessed. Models were adjusted as previously described and included age, study site, enrollment period, and self-reported race/ethnicity [6]. We performed stratified analyses by 1) time of illness onset using February – May 2021 as a pre-Delta variant period and July – September 2021 as the Delta variant-predominant period and 2) by age group (as 12-49 years and >49 years due to sample size) [10].

We performed several sensitivity analyses. We compared VE using plausible self-report of vaccination to VE using only documented vaccination status, where plausibility was determined by ability to report credible location of vaccination, as previously described [6]. We also compared findings when persons who identified as working in healthcare were excluded and also when persons with unknown status were classified as having had no known contact. Statistical analyses were conducted using SAS version 9.4 (SAS Institute Inc., Cary, NC, USA). This research activity involving human subjects was reviewed by the Institutional Review Boards (IRB) of the Centers for Disease Control and Prevention (CDC) and Baylor Scott and White Health, Marshfield Clinic Research Institute, University of Michigan, Henry Ford Health System, and University of Pittsburgh and was conducted consistent with applicable federal law and CDC policy (See 45 C.F.R. part 46; 21 C.F.R. part 56). The Kaiser Permanente Washington Health Research Institute IRB determined this activity was research not involving human subjects under a nondisclosure agreement with CDC.

## Results

Of 3,384 symptomatic persons aged ≥12 years enrolled from February 1 through September 30, 2021, information on contact with a person with confirmed COVID-19 disease during the 14 days before illness onset was available from 2,229 (65.9%) participants: 451 (20%) reported contact and 1,778 (80%) reported no known contact. Among participants reporting contact, 283 (63%) were SARS-CoV-2 test-positive cases compared with 331 (19%) of participants without known contact. Participants reporting contact with persons with COVID-19 disease were more likely than those with unknown contact to be healthcare workers, aged <65 years, household contacts of children aged <12 years, tested for SARS-CoV-2 less than four days since illness onset, and have no documented COVID-19 vaccination (Table 1).

**Table 1.** Characteristics of enrolled participants by self-reported known contact with a person with SARS-CoV-2 in the 14 days prior to illness onset, U.S. Flu VE Network, February 1– September 30, 2021

|  |  | **Known Contact** |  | **No Known Contact** |  |  |
| --- | --- | --- | --- | --- | --- | --- |
|  | **Total** | **n** | **col %** | **n** | **col %** | **p-value<sup>a</sup>** |
| **Total** | 2229 | 451 | 100 | 1778 | 100 |  |
| **Age Group (years)** |  |  |  |  |  | <0.01 |
| 12–15 | 99 | 18 | 4 | 81 | 5 |  |
| 16–49 | 1182 | 267 | 59 | 915 | 51 |  |
| 50–64 | 523 | 112 | 25 | 411 | 23 |  |
| ≥65 | 425 | 54 | 12 | 371 | 21 |  |
| **Study Site** |  |  |  |  |  | <0.01 |
| Michigan | 312 | 102 | 23 | 210 | 12 |  |
| Pennsylvania | 358 | 75 | 17 | 283 | 16 |  |
| Texas | 377 | 102 | 23 | 275 | 15 |  |
| Washington | 740 | 90 | 20 | 650 | 37 |  |
| Wisconsin | 442 | 82 | 18 | 360 | 20 |  |
| **Sex** |  |  |  |  |  | 0.45 |
| Female | 1438 | 298 | 66 | 1140 | 64 |  |
| Male | 790 | 153 | 34 | 637 | 36 |  |
| **Race/Ethnicity** |  |  |  |  |  | 0.07 |
| Black Non-Hispanic | 186 | 33 | 7 | 153 | 9 |  |
| Hispanic | 159 | 38 | 9 | 121 | 7 |  |
| Other Non-Hispanic | 211 | 30 | 7 | 181 | 10 |  |
| White Non-Hispanic | 1661 | 346 | 77 | 1315 | 74 |  |
| **Underlying Condition<sup>b</sup>** |  |  |  |  |  | 0.12 |
| No | 1402 | 270 | 61 | 1132 | 65 |  |
| Yes | 789 | 174 | 39 | 615 | 35 |  |
| **Child aged <12 years living in household** |  |  |  |  |  | <0.01 |
| None | 1589 | 286 | 63 | 1303 | 73 |  |
| ≥1 | 640 | 165 | 37 | 475 | 27 |  |
| **Healthcare Worker<sup>c</sup>** |  |  |  |  |  | <0.01 |
| No | 1770 | 329 | 79 | 1441 | 88 |  |
| Yes | 275 | 87 | 21 | 188 | 12 |  |
| **Documented 2020-21 influenza vaccination** |  |  |  |  |  | <0.01 |
| No | 1606 | 368 | 82 | 1238 | 70 |  |
| Yes | 623 | 83 | 18 | 540 | 30 |  |
| **Prior SARS-CoV-2 Infection<sup>d</sup>** |  |  |  |  |  | 0.87 |
| No | 1993 | 405 | 91 | 1588 | 91 |  |
| Yes | 192 | 40 | 9 | 152 | 9 |  |
| **Symptom Onset to Specimen Collection Interval (days)** |  |  |  |  |  | <0.01 |
| 0–3 | 1134 | 258 | 57 | 876 | 49 |  |
| 4–6 | 771 | 128 | 28 | 643 | 36 |  |
| 7–10 | 324 | 65 | 14 | 259 | 15 |  |
| **COVID-19 Vaccination Status<sup>e</sup>** |  |  |  |  |  | <0.01 |
| Fully vaccinated | 1134 | 183 | 43 | 951 | 56 |  |
| Partially vaccinated | 154 | 26 | 6 | 128 | 8 |  |
| Unvaccinated | 828 | 220 | 51 | 608 | 36 |  |
| **SARS-CoV-2 Status** |  |  |  |  |  | <0.01 |
| Negative | 1615 | 168 | 37 | 1447 | 81 |  |
| Positive | 614 | 283 | 63 | 331 | 19 |  |

<sup>a</sup> P-value is for the chi-square test where $p < 0.05$ was considered statistically significant
<sup>b</sup> Underlying condition (e.g., heart disease, lung disease, diabetes, cancer, liver or kidney disease, immune suppression, or high blood pressure) is self-reported; 38 participants (7 with known contact and 31 participants with no known contact) missing underlying condition status
<sup>c</sup> Work in healthcare setting is self-reported; 21 participants missing healthcare worker status; 163 participants <18 years old were not asked this question
<sup>d</sup> Prior SARS-CoV-2 infection is self-reported; 44 participants (6 with known contact and 44 with no known contact) missing prior infection status
<sup>e</sup> There were 112 participants who had indeterminate vaccination status (22 with known contact and 90 with no known contact)

Among participants reporting known contact, 183 (43%) were fully vaccinated ≥14 days before illness onset versus 951 (56%) of those without known contact (Table 1). Most of the 1,288 vaccinated participants received mRNA vaccines (1,204, 93%); 761(59%) received Pfizer-BioNTech and 443 (34%) received Moderna. Few participants (81, 6%) received Johnson and Johnson/Janssen vaccine, or a vaccine of unknown type (3, <1%). Over 90% of vaccinated participants were vaccinated prior to mid-May 2021. The median number of days between most recent dose and onset of symptoms was 113 days (interquartile range 67-155) with no differences observed by the product received.

Among all participants with non-indeterminate vaccination status (n=2,116), VE of documented full vaccination against laboratory-confirmed, symptomatic COVID-19 disease with any COVID-19 vaccine was 72% (95% Confidence Interval [CI]: 64-78). Similar estimates were observed among participants with and without known contact (Figure 1). Adjusted VE among fully vaccinated participants with known contact was 71% (95% CI:49-83) compared to 80% (95% CI: 72-86) among fully vaccinated participants with no known contact (p-value for interaction = 0.2). Significant interaction was not observed for any comparison (p-value ≥0.1 for all). In sensitivity analyses, overall VE was similar when known healthcare workers were excluded, participants with unknown contact (n=1,155) were classified as “no known contact,” plausible self-reported doses were incorporated, and partially vaccinated participants were included (Supplemental Table).

**Figure.**
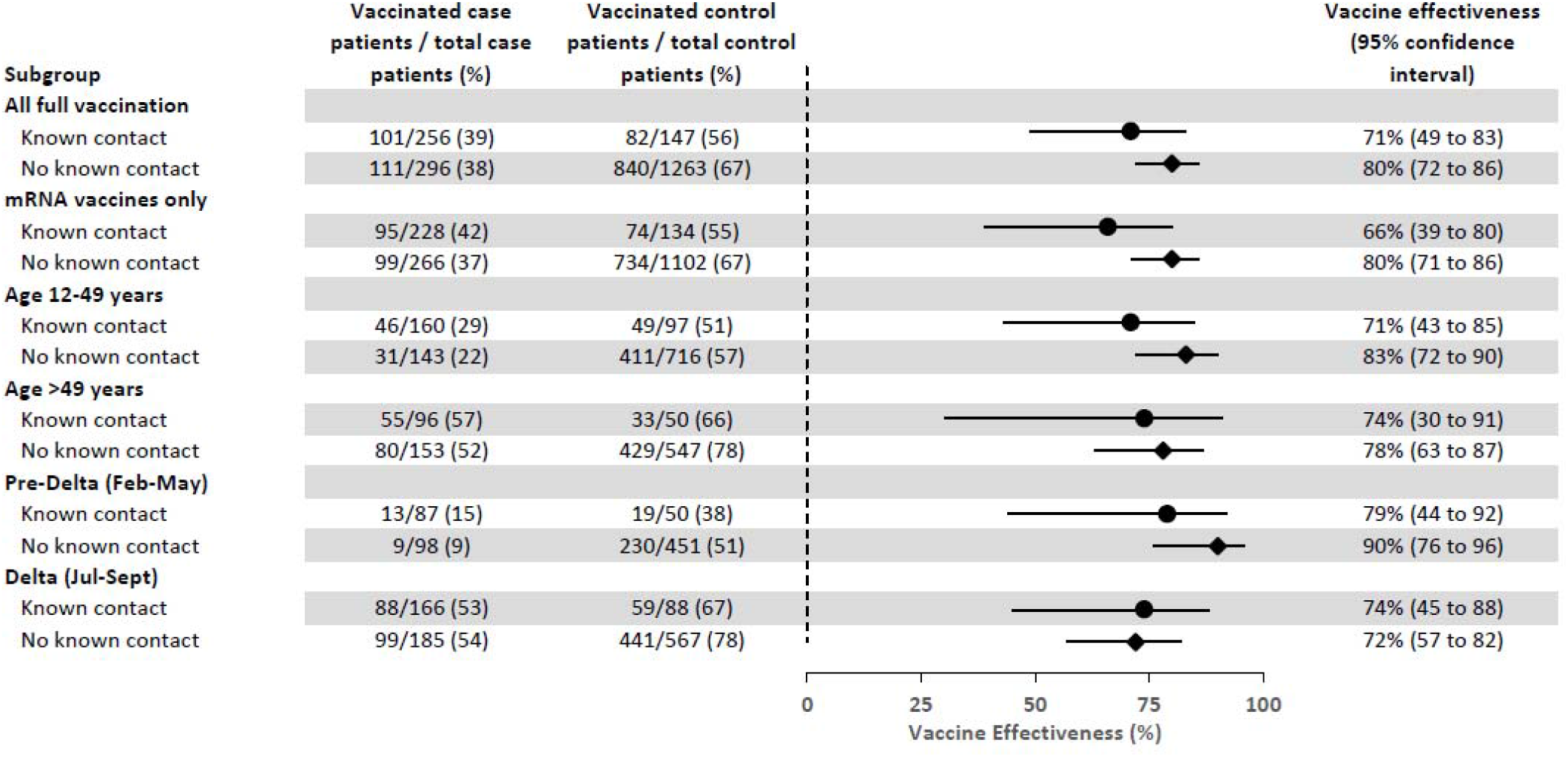
Estimates of vaccine effectiveness^a^ against laboratory-confirmed symptomatic COVID-19 among participants with and without reported known contact with persons with COVID-19 disease during 14 days before illness onset, US Flu VE Network, February 1–September 30, 2021. CI=Confidence interval ^a^ Vaccine effectiveness (VE) of full vaccination by documented records versus unvaccinated. Model adjusted for study site, age in years (continuous), enrollment period (natural cubic spline with 3 percentile knots of interval between January 1, 2021 and illness onset date), and self-reported race/ethnicity.

VE point estimates of full vaccination against symptomatic laboratory-confirmed COVID-19 disease among all participants regardless of reported exposure status differed by mRNA product [Moderna VE 81% (95% CI: 73-86), Pfizer VE 66% (95% CI: 56-73)]; age group [70% (95% CI: 59-78) among participants aged 16-49 years, 80% (95% CI: 68-88) among participants 50-64 years, and 61% (95% CI: 29-79) among participants aged ≥65 years]; and enrollment period [86% (95% CI 77-92) for participants with onset in the pre-Delta-variant period and 59% (95% CI: 45-69) for participants with onset during Delta variant-predominance period].

## Discussion

Vaccination against COVID-19 disease provided protection against laboratory-confirmed, symptomatic outpatient illness among individuals who reported known contact with a person with COVID-19 disease. Participants who reported known contact with a person with COVID-19 disease were more likely to test positive for SARS-CoV-2 infection compared to participants who reported no known contact. Those who reported a known contact were more likely to report living in a household with at least one child aged <12 years, or to report work in a healthcare setting; our findings were robust when persons who work in a healthcare setting were excluded. In addition, we did not detect a statistically significant difference by known contact status when participants were further stratified by illness onset into pre-Delta variant versus Delta variant circulation periods.

Having contact with a known person with SARS-CoV-2 infection substantially increases the likelihood of testing positive for SARS-CoV-2. Contact tracing and transmission studies suggest that household settings have the highest secondary attack rates, with an estimated pooled secondary transmission rate of 21.1% (95% CI: 17.4-24.8) [3]. Although we were unable to categorize the setting of known exposure in our study, other studies have compared secondary transmission from a household contact compared to other forms of contact and highlighted the importance of household transmission compared to occupational, social, or transportation exposures [11-13]. The importance of household transmission is likely due to prolonged exposures in close proximity with fewer protective measures in place. In one study, unvaccinated or partially vaccinated persons were more likely to transmit SARS-CoV-2 virus compared to fully vaccinated persons [13]. We show equivalent, high levels of protection of full vaccination against symptomatic, laboratory-confirmed COVID-19 disease regardless of known contact.

We build upon prior published findings from the Flu VE Network in several ways. This analysis includes four additional months of data compared to an earlier evaluation of COVID-19 VE between February and May 2021 [6]. Despite predominance of the Delta variant in the latter study period [10], our findings show protection against laboratory-confirmed symptomatic illness. A decline in VE point estimates in the latter study period could be attributed to reduced protection against the Delta or a result of waning protection of the initial vaccination series [14, 15]. However, this study was underpowered to evaluate and disentangle these factors. Similar to other published reports, we detected a lower adjusted VE of Pfizer-BioNTech vaccine when compared to Moderna vaccine [16]. This analysis summarizes VE of full vaccination against laboratory-confirmed symptomatic outpatient illness in the period of time prior to when third (i.e., booster) doses became available and were recommended, as well as the period of time when vaccination was recommended for US children aged five to twelve years of age [17, 18]. Booster doses of all COVID-19 vaccines are currently available for adults aged ≥18 years in the US [19].

Our study was subject to at least five limitations. First, we did not assess the nature of reported contact and do not have information about whether the exposure was a household member, occupational, or other type of exposure. Second, we did not collect information about the timing of the reported known contact within the 14 days prior to illness onset. Participants could have been infected prior to the reported exposure. Third, some participants might have been aware of their SARS-CoV-2 test status when they completed the enrollment questionnaire, which could have influenced responses to the known contact question [11]. While the test-negative design reduces bias due to differences in healthcare-seeking behavior among vaccinated and unvaccinated persons [9], vaccinated cases could have been more motivated to participate in this study. Fourth, we did not ask about NPIs or duration of contact of the known exposure. Differences in exposures and prevention measures among vaccinated and unvaccinated participants could have been associated with likelihood of testing positive for SARS-CoV-2 infection. Finally, our study was unable to account for differences in timing of vaccination.

This study contributes to growing evidence of COVID-19 VE against symptomatic illness, including sustained protection in the Delta-variant period, among members of the general population who have contact with persons with COVID-19 disease. These findings support recommendations for COVID-19 vaccination for the prevention of symptomatic illness and highlight the importance of continued efforts to increase vaccination coverage.

## Data Availability

De-identified dataset can be made available upon request

## Acknowledgements

Hannah Berger, Joshua Blake, Keegan Brighton, Gina Burbey, Deanna Cole, Linda Heeren, Erin Higdon, Lynn Ivacic, Julie Karl, Sarah Kopitzke, Erik Kronholm, Jennifer Meece, Nidhi Mehta, Vicki Moon, Cory Pike, Carla Rottscheit, Jackie Salzwedel, Marshfield Clinic Research Institute, Marshfield, Wisconsin; Alanna Peterson, Linda Haynes, Erin Bowser, Louise Taylor, Karen Clarke, Christopher Deluca, Todd M. Bear, Klancie Dauer, Goundappa K Balasubramani, Robert Hickey, Monika Johnson, Donald B. Middleton, Jonathan M. Raviotta, Theresa Sax, Miles Stiegler, Michael Susick, Joe Suyama, Rachel Taber, Alexandra Weissman, John V. Williams, University of Pittsburgh Schools of the Health Sciences and University of Pittsburgh Medical Center, Pittsburgh, Pennsylvania; Adam Lauring, Joshua G. Petrie, Lois E. Lamerato, E.J. McSpadden, Caroline K.Cheng, Rachel Truscon, Samantha Harrison, Armanda Kimberly, Anne Kaniclides, Kim Beney, Sarah Bauer, Michelle Groesbeck, Joelle Baxter, Rebecca Fong, Drew Edwards, Weronika Damek Valvano, Micah Wildes, Regina Lehmann-Wandell, Caitlyn Fisher, Luis Gago, Marco Ciavaglia, Kristen Henson, Kim Jermanus, Alexis Paul, University of Michigan, Ann Arbor, and Henry Ford Health System, Detroit, Michigan; Eric Hoffman, Martha Zayed, Marcus Volz, Kimberly Walker, Arundhati Rao, Manohar Mutnal, Michael Reis, Lydia Requenez, Amanda McKillop, Spencer Rose, Kempapura Murthy, Chandni Raiyani, Natalie Settele, Jason Ettlinger, Courtney Shaver, Elisa Priest, Jennifer Thomas, Alejandro Arroliga, Madhava Beeram, Baylor Scott & White Health, Temple Texas; C. Hallie Phillips, Erika Kiniry, Stacie Wellwood, Brianna Wickersham, Matt Nguyen, Rachael Burganowski, Suzie Park, Kaiser Permanente Washington Research Institute, Seattle, Washington

## Disclaimers

The findings and conclusions in this report are those of the authors and do not necessarily represent the official position of the Centers for Disease Control and Prevention / Agency for Toxic Substances and Disease Registry.

Vaccination data from Pennsylvania were supplied by the Bureau of Health Statistics & Registries, Pennsylvania Department of Health, Harrisburg, Pennsylvania. The Pennsylvania Department of Health specifically disclaims responsibility for any analyses, interpretations, or conclusions.

---

**Supplemental Table.** Results of sensitivity analyses of vaccine effectiveness against laboratory-confirmed symptomatic COVID-19, US Flu VE Network, February 1–September 30, 2021

|  | SARS-CoV-2-Positive CLI<br>(Cases) |  |  | SARS-CoV-2-Negative CLI<br>Controls |  |  | Adjusted* VE |  |
| --- | --- | --- | --- | --- | --- | --- | --- | --- |
|  | #<br>Vaccinated | Total | %<br>Vaccinated | #<br>Vaccinated | Total | %<br>Vaccinated | VE % | (95% CI) |
| **Include partially vaccinated participants** |  |  |  |  |  |  |  |  |
| Known contact | 117 | 268 | 44 | 98 | 161 | 61 | 73 | (54 to 84) |
| No known contact | 133 | 309 | 43 | 960 | 1355 | 71 | 78 | (69 to 84) |
| **Exclude healthcare workers** |  |  |  |  |  |  |  |  |
| Known contact | 76 | 215 | 35 | 58 | 110 | 53 | 73 | (47 to 86) |
| No known contact | 101 | 267 | 38 | 738 | 1116 | 66 | 80 | (70 to 86) |
| **Assume persons with unknown status had no known contact** |  |  |  |  |  |  |  |  |
| No known contact | 230 | 542 | 42 | 977 | 1486 | 66 | 71 | (62 to 78) |
| **Include plausible self-reported doses** |  |  |  |  |  |  |  |  |
| Known contact | 104 | 255 | 41 | 84 | 147 | 57 | 72 | (51 to 84) |
| No known contact | 117 | 293 | 40 | 846 | 1241 | 68 | 80 | (71 to 86) |

CLI = COVID-19-like illness
\*Vaccine effectiveness (VE) adjusted for study site, age in years (continuous), enrollment period (natural cubic spline with 3 percentile knots of interval between January 1, 2021 and illness onset date), and self-reported race/ethnicity.

